# Whole-genome sequencing of 197 cases with Parkinson’s Disease reveals novel pathogenic variants in the Indian population

**DOI:** 10.1101/2025.10.19.25338055

**Authors:** Hiba Ali, Banashree Mondal, Andie Pinto, Camellia Sarkar, Bikram Kumar Panda, Khader Valli Rupanagudi, Vikram V Holla, Tarunya Nagaraj, Shubha GS Bhat, Shweta Prasad, Mahima Bhardwaj, Pooja Mailankody, Rohan R Mahale, Nitish Kamble, Ravi Yadav, Pramod Kumar Pal, Albert Stezin, Latha Diwakar, Shweta Ramdas

## Abstract

**Background:** Most genetic studies of Parkinson’s disease (PD) have been performed in European cohorts, and large-scale studies in Indian populations are lacking. Consequently, many variants common in Europeans are rare or absent in Indian patients, emphasizing the need for population-specific genomic studies.

**Objectives:** To identify novel genetic variants contributing to PD in Indian populations, and quantify the polygenic burden of the disease in cases versus controls.

**Methods:** We performed WGS of 197 cases (106 early-onset PD (EOPD) and 91 late-onset PD (LOPD)) and 32 controls from India. We prioritized pathogenic variants in 67 PD-associated genes using ACMG guidelines. We quantified polygenic risk scores (PRS) for PD in our cohort using summary statistics from European GWAS, and tested PRS differences between cases and controls.

**Results:** We identified likely pathogenic variants in 31 cases across eight PD-associated genes (*GBA1, PRKN, PINK1, PARK7, SYNJ1, SNCA,* and *PLA2G6*), giving a diagnostic yield of 27.63% in EOPD and 2.19% in LOPD. Of 29 identified variants, seven were novel. *GBA1* was the most frequently mutated gene, followed by *PRKN*. Genes with pathogenic variants were enriched in lysosomal pathways. Cases had significantly higher PRS than controls, confirming a polygenic contribution to PD risk.

**Conclusions:** This study represents one of the first WGS-based genetic investigations of PD in India with both EOPD and LOPD cases. Novel variants in *PRKN, GBA1, LRRK2, ATXN2* and *SNCA* point to the presence of population-specific variants, and reinforce the importance of genetic studies in diverse populations.

## Introduction

Parkinson’s disease (PD) is a complex neurodegenerative disorder clinically characterized by bradykinesia, rigidity, and resting tremors along with non-motor symptoms such as neuropsychiatric involvement, autonomic dysfunction, sleep disturbances, and olfactory loss^1^. The pathological hallmarks of PD include dopaminergic neuronal loss in the substantia nigra, and intracellular accumulation of α-synuclein aggregates known as Lewy bodies^2^. The global burden of PD has increased substantially in recent decades, with more than six million people affected in 2015 and projections suggesting over 12 million cases by 2040^3^. This steep rise, driven by aging populations and improved diagnostics, has led some to describe PD as a “pandemic”^4^. The mounting public health burden highlights an urgent need to understand its etiology across ancestrally and geographically diverse populations.

Genetic studies have greatly advanced our understanding of PD, with discoveries of both monogenic forms and polygenic contributions to disease risk^5^. 10-15% of PD cases have a family history, with 5-10% showing a clear Mendelian inheritance pattern; most cases (85-90%) are sporadic^6^. The most common monogenic PD-related genes are *SNCA, LRRK2, PRKN (PARK2), PINK1, DJ1, GBA1*^7–10^. Mutations in *GBA1* and *LRRK2* are among the most frequent risk variants worldwide (1-10% in different cohorts), with some ethnicity-specific hotspots^10–12^. Apart from Mendelian variants, many common variants of small effect contribute to PD, as revealed by genome-wide association studies (GWAS)^5,11^. While rare mutations in genes like *SNCA*, *LRRK2*, and *PRKN* cause familial PD^13^, recent meta-analyses have identified 90 risk loci explaining 20–30% of risk^14^. These findings have led to the development of polygenic risk scores (PRS), which can stratify individuals by their genetic risk. Apart from the genetic risk, environmental influences like pesticide exposure, metals, solvents, and even lifestyle factors affect PD risk^15–18^.

Until recently, most genetic studies have focused on individuals of European ancestry^19^. Over 80% of PD GWAS participants are of European descent^20^, and there are few large-scale studies in Indian cohorts^10^. Many variants identified in European cohorts are rare or absent in Indian populations^21^. For example, *LRRK2* p.G2019Ss, common in Europeans and Ashkenazi Jews, is infrequent in Indian PD patients^21,22^. Moreover, most PRS models are based on European datasets and may perform poorly in other populations due to differences in allele frequency, linkage disequilibrium and variant effect sizes^23^. This highlights the need for population-specific PRS development to improve risk prediction and enable more inclusive precision medicine approaches.

Existing Indian PD studies are limited by moderate sample sizes, a focus on known candidate genes, or reliance on SNP-array or exome-based approaches^24,25^. These methods primarily detect common coding variants and often miss structural variants (SVs), deep intronic mutations, and regulatory changes that may contribute to disease risk. Recent discoveries highlight the role of such variant classes in PD pathogenesis. For instance, an intronic enhancer variant in *SNCA* ^26^ and cryptic splice site mutations in *PARK7*^27,28^ impact PD risk by affecting gene function.

In this study, we performed whole-genome sequencing (WGS) of 197 Indians with PD to map its genetic landscape in this underrepresented population. WGS offers a comprehensive platform for capturing the full spectrum of genomic variation across coding and non-coding regions, including rare single nucleotide variants (SNVs), SVs, repeat expansions, and splice-altering changes^29,30^. We aimed to uncover population-specific variation and novel loci, advancing understanding of PD genetic architecture in India.

## Methods

### Cohort description

We enrolled 229 individuals in our cohort from the Movement Disorders Clinic at the National Institute of Mental Health and Neurosciences (NIMHANS), a tertiary referral hospital in India, including 197 PD patients from across India. 106 individuals had early-onset PD (EOPD), defined as age of onset (AAO) < 50 years, and 91 had late-onset PD (LOPD), with an AAO greater than 50 years. The average AAO was 38.95 ± 7.92 years for EOPD and 57.62 ± 5.54 years for LOPD. 72 out of 106 EOPD cases (67.9%), and 71 out of 91 LOPD cases (78%) were male. The dataset also included 32 healthy controls (HC). Table S1 contains demographic information for all 229 samples. Principal component analysis of genetic data showed clustering of individuals by state (Figure S1).

### Participant selection criteria

Diagnoses of PD was based on the International Parkinson and Movement Disorders Society (MDS) clinical diagnostic criteria for PD^31^ applied by the trained neurologists (PKP, RY, RM, NK, PM, and VVH). Between 2022 and 2024, all patients diagnosed with PD during routine clinical evaluation were offered participation in this study. Demographic and clinical details were collected during a structured interview which also included details of family history of PD, age at onset, and clinical details such as motor and non-motor symptoms. A small number of HC were carefully age- and sex-matched to PD cases and recruited from the same geographic and ethnic background to minimize population stratification. Individuals with a personal or family history of PD or other neurodegenerative conditions were excluded from the HC group.

### Ethical considerations

Institutional Ethics Committee approval from NIMHANS and the Centre for Brain Research was obtained prior to the study. All participants provided written informed consent, acknowledging voluntary participation, and the use of their anonymized genetic data for research purposes. All sample identifiers were generated internally within the research group and were never known to study participants.

### Genomic DNA processing

We collected whole blood from the study participants and extracted genomic DNA using standard protocols (QIAamp DNA Blood Midi Kit, Qiagen). We measured DNA concentration using the Qubit fluorometer (Thermo Fisher Scientific, USA). We constructed DNA libraries using TruSeq DNA PCR-free kit from Illumina. Samples were sequenced on the Illumina NovaSeq 6000 using 150 bp paired-end reads, achieving an average coverage of 54X. We converted raw BCL files to FASTQ format using bcl2fastq2 v2.20 (Illumina, Inc.). We assessed read quality using FastQC v0.12.1^32^ and MultiQC v1.31^33^, and trimmed adapters and low-quality bases using Trimmomatic v0.39^34^.

### Variant calling and interpretation

We aligned sequencing reads to the GRCh38 reference genome without alt contigs using BWA-MEM2 v2.2.1^35^. We followed Genome Analysis Toolkit (GATK) v4.4.0 best practices^36^ for variant calling and preprocessing, including duplicate marking, base quality score recalibration, joint genotyping, and variant quality filtering. We annotated variants using SnpEff^37^ and ANNOVAR^38^ with reference databases including gnomAD^39^. For splice variants, we used SpliceAI^40^ score >0.8 to filter potential splice disrupting splice variants. We defined rare variants as those with a minor allele frequency (MAF) < 1% in gnomAD to classify them as pathogenic (P), likely pathogenic (LP), variants of uncertain significance (VUS), or benign as per the American College of Medical Genetics and Genomics (ACMG) guidelines^41^.

### SVs calling and prioritization

We called SVs using five tools: CNVnator^42^ (v0.4.1), LUMPY^43^ (v0.3.1), Manta^44^ (v1.6.0), Delly^45^ (v1.2.6), and BreakDancer^46^ (v1.4.5). For insertions, we used Manta, BreakDancer, INSurVeyor^47^ (v1.1.3) and Delly, since CNVnator and LUMPY don’t call insertions. We analysed deletions, insertions, duplications, and inversions considering only variants with FILTER=PASS. VCFs for each sample were merged using SURVIVOR^48^ (v1.0.3) with the following parameters: maximum distance between breakpoints = 1000 bp, minimum support = 1 caller, require same SV type and strand, enable distance estimation, and a minimum SV size of 50 bp. We prioritized high-confidence variants supported by all callers (SUPP=4 for insertions, and SUPP=5 for all other classes) that overlapped exons in any of the 67 PD-associated genes. Final candidate variants were assessed for pathogenicity according to ACMG criteria and visually validated using the Integrative Genomics Viewer (IGV, v2.16.2)^49^.

### Polygenic risk score (PRS)

We estimated PRS for our dataset using LDpred-2 v1.11.6^50^. We used summary statistics from the largest European-ancestry PD GWAS as the base dataset^14^. To ensure high-quality variant data, SNPs with a genotype missing rate >1% were excluded along with individuals with >1% missing genotypes. We filtered out SNPs that significantly deviated from Hardy-Weinberg equilibrium (p < 1×10⁻⁶) and with MAF < 1%. To reduce redundancy due to linkage disequilibrium, we performed LD pruning using a sliding window of 200 SNPs, moving 50 SNPs at a time, and retained variants with pairwise r² values below 0.25^51^.

Common SNPs filtered from the two datasets were restricted to those present in HapMap3+reference panel to ensure high variant inclusion. Finally, we computed LD matrix which along with the base data was used as input for the LDpred2-auto model^52^. Since our dataset has very few controls, we used 1,163 samples from the GenomeAsia project^53^ to assess PRS in an external ancestry-matched control population. These controls were recruited from outpatient clinics and population-based studies across multiple cities in India, and were genotyped using the Illumina Global Screening Array, which includes over 600,000 genome-wide markers^54^. The distributions of PRS were compared between cases and controls as well as among PD subgroups categorized based on the presence of pathogenic mutations.

### Pathway enrichment analysis

We performed geneset enrichment analysis to investigate the biological relevance of rare variants in PD cases. We used Enrichr^55^ to test enrichment against the KEGG_2021_Human, Reactome, and Gene Ontology (GO) Biological Process databases. Pathways with an adjusted *P*-value < 0.05 (Benjamini-Hochberg false discovery rate) were considered significantly enriched.

### Gene burden analysis

To identify genes with an excess burden of rare variants associated with PD, we used the consistent summary counts based rare variant burden test (CoCoRV) framework^56^. We compared our case cohort against gnomAD public summary data, filtering DP ≥ 10, GQ ≥ 20 and excluding genomic regions flagged as problematic in gnomAD (segmental duplications, low-complexity regions, and other blacklisted sites based on gnomAD QC). Cases were assigned to gnomAD populations using a pre-trained random forest classifier to account for ancestry differences. We limited analysis to regions with ≥10X coverage in ≥90% of samples and only retained loss-of-function and missense variants with REVEL ≥ 0.65. Burden testing used a dominant genetic model with MAF ≤ 1e-2, while accounting for linkage disequilibrium. False discovery rate was controlled using a resampling-based approach, and potential residual confounding was evaluated using genomic inflation factor (λ).

## Results

### Prioritization of protein-coding and splicing SNVs and indels

We called genotypes for 229 samples at 29,932,165 variants using the GATK best practice variant calling pipeline. We filtered out common variants with >1% MAF in gnomAD v4, giving us 18,259,705 variants (61%). Among these, 131,141 (0.72%) were missense, frameshift, stop-gained or splice variants. 435 rare variants were in 67 genes previously implicated in PD (Table S2). We removed variants found in more than one control, and those for which carrier genotypes didn’t match the known pattern of inheritance for the gene. These criteria filtered our list to 166 variants. We further prioritized these variants using ACMG guidelines.

23 cases (Figure 1A, 1B; Table 1, S3) carried pathogenic or likely-pathogenic (P/LP) variants in eight PD-associated genes: *GBA1, PRKN, PINK1, PARK7, SYNJ1, SNCA* and *PLA2G6*. Ten out of these 23 cases (all EOPD) were homozygous or compound heterozygous for the identified variants. The dominance of homozygous mutations in EOPD is consistent with patterns of monogenic early-onset disease^56–58^.

**Figure 1.**
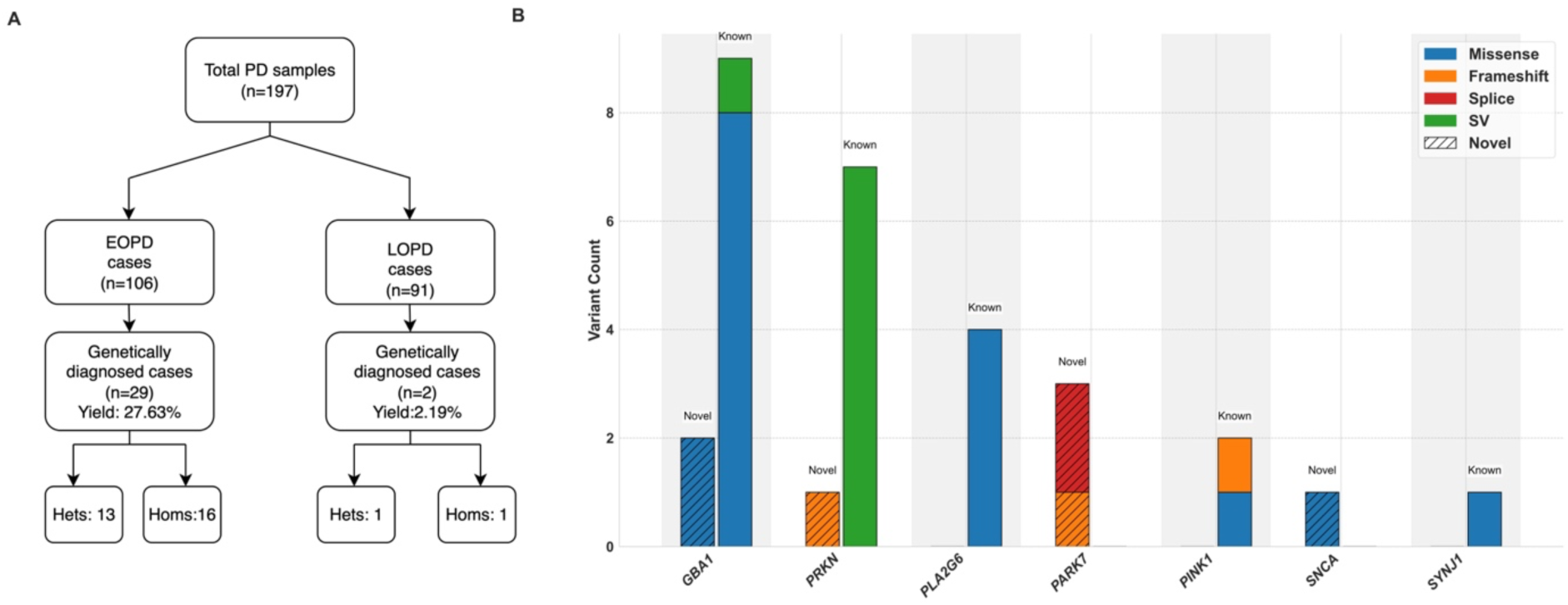
Genetic diagnostic yield and pathogenic variant distribution across PD-associated genes. **(A)** illustrates the diagnostic yield, and the number of heterozygous (hets) and homozygous (homs) carriers in EOPD and LOPD cases. **(B)** shows the distribution of pathogenic mutations across the analysed PD-associated genes

**Figure 2.**
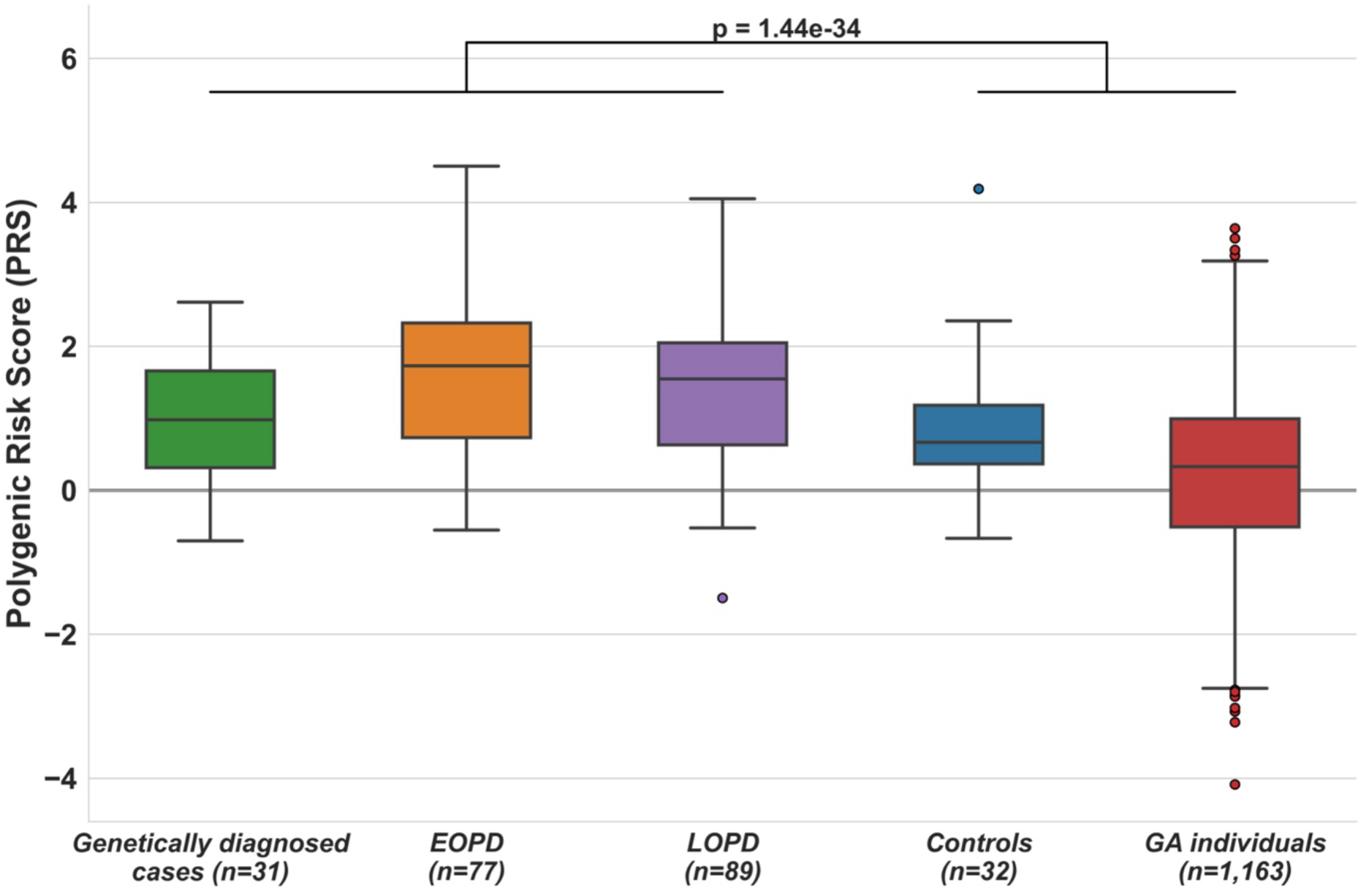
Distribution of PRS in cases, cases without a Mendelian variant, and controls. Distribution of Polygenic Risk Scores (PRS) values among five groups: genetically diagnosed cases, individuals with EOPD, individuals with LOPD), healthy controls (HC), and GenomeAsia individuals (GA)

**Table 1.** Pathogenic and likely pathogenic variants identified in 23 PD cases. Each entry includes the sample id, disease subtype, and family history (F: familial, S: sporadic), the affected gene and zygosity status (het: heterozygous, hom: homozygous, compound het: compound heterozygous), and type of mutation (missense, frameshift, or splice-site). Variants are also classified as *novel* (not previously reported in PD or reference databases).

| Sample ID | Disease subtype/<br>family history | Variant ID | Gene | Zygosity | Type |
| --- | --- | --- | --- | --- | --- |
| YLOP D177 | EOPD/S | c.90+1delG [novel] | *PARK7* | *hom* | splice |
| YLOP D185 | EOPD/F | c.90+1delG [novel] | *PARK7* | *hom* | splice |
| YLOP D-37 | EOPD/S | c.456delT [novel] | *PARK7* | *hom* | frame shift |
| YLOP D-02 | LOPD/S | c.776A>T [novel] | *GBA1* | *het* | missense |
| YLOP D_96 | EOPD/F | c.711G>C [novel] | *GBA1* | *het* | missense |
| YLOP D139 | EOPD/S | c.711G>C [novel] | *SNCA* | *het* | missense |
| YLOP D28 | EOPD/F | c.374dupC [novel] | *PRKN* | *hom* | frame shift |
| YLOP D211 | EOPD/S | c.85_106delTACGGCTTG<br>GGGCGGCCCGGGCC | *PINK1* | *hom* | frame shift |
| YLOP D157 | EOPD/S | c.1153T>C | *PINK1* | *hom* | missense |
| YLOP D_62 | EOPD/S | c.1448T>G | *GBA1* | *het* | missense |
| YLOP D128 | EOPD/S | c.1448T>G | *GBA1* | *het* | missense |
| YLOP D29 | EOPD/S | c.1448T>C | *GBA1* | *het* | missense |
| YLOP D184 | EOPD/S | c.1448T>C | *GBA1* | *het* | missense |
| YLOP D152 | EOPD/S | c.1342G>C | *GBA1* | *het* | missense |
| YLOP D193 | EOPD/S | c.1342G>C | *GBA1* | *het* | missense |
| YLOP D227 | EOPD/S | c.827C>T | *GBA1* | *het* | missense |
| YLOP D209 | EOPD/S | c.667T>C | *GBA1* | *het* | missense |
| YLOP D208 | EOPD/S | c.509G>A | *GBA1* | *hom* | misse<br>nse |
| YLOP D40 | EOPD/S | c.492C>G | *GBA1* | *het* | misse<br>nse |
| YLOP D 20 | EOPD/S | c.44T>C | *GBA1* | *het* | misse<br>nse |
| YLOP D_90 | EOPD/F | c.773G>A | *SYNJ1* | *hom* | misse<br>nse |
| YLOP D238 | EOPD/S | c.773G>A; c.2239C>T | *PLA2 G6* | *compound het* | misse<br>nse |
| YLOP D50 | EOPD/F | c.2239C>T;<br>c.1898C>T | *PLA2 G6* | *compound het* | misse<br>nse |

Figure 1B shows the distribution of pathogenic variants (SNVs, indels, and SVs) found in PD-associated genes. *GBA1* was the most commonly mutated gene in our cohort, with 13 cases carrying P/LP variants. These included two variants previously not reported for PD pathogenesis, one of which was absent from gnomAD. This is unlike a few other genomic studies in Indian PD cohorts, which did not report a high number of *GBA1*^59^. Apart from the mutations in these 23 diagnosed cases, we also screened for mutations in 37 genes associated with parkinsonism, dystonia, and ataxia. Several additional variants were identified that may be relevant to disease etiology but currently lack sufficient evidence (in terms of zygosity or direct relevance to the phenotype) to be classified as causal. Two cases carried a heterozygous novel pathogenic *ATXN2* mutation. Eleven cases carried heterozygous pathogenic variants in genes primarily associated with recessive parkinsonism (*DNAJC6, VPS13C, SPG11* (two cases), *FBXO7, TH* (two cases), *CP, MYORG* (two cases), and *AOPEP)* (Table S5). The list of all variants in 37 genes is listed in Table S6.

We manually annotated and curated the list of VUS in the 67 PD-related genes to prioritize variants in this list (Table S4), and to identify potential likely-pathogenic variants that were missed by previous annotation methods. One LOPD case carried a novel, highly damaging variant in *LRRK2* (pAla1442Pro), absent from gnomAD. While the VUS status for this variant is based on its unknown effect on protein structure, AlphaMissense^61^ predicts a strong effect on protein structure (pathogenicity score of 0.987), leading us to prioritize this as a likely causal variant that should be followed up in functional studies. Genes with rare VUS (Table S7) seen only in PD cases were enriched in pathways of neurodegeneration (Figure S4A) (odds ratio 4.4; adjusted *P* = 1.26e-05), suggesting that this list of variants and genes includes truly pathogenic variants that are relevant for PD in this cohort. Genes with P/LP (Table S8) variants in were also enriched in PD (odds ratio 7.25; *P =* 2.9e-04) and lysosome pathways (odds ratio 14.53; *P =* 7.32e-06; Figure S4D, Table S14).

### Pathogenic SVs in PD

We called SVs using five different SV callers, prioritizing those called by all five to minimize false positive calls. We specifically examined SVs overlapping exons of 67 PD-associated genes that are found in cases and no more than 1 control. After filtering for PD genes and inheritance pattern of the gene, no insertions or duplications were prioritized. Seven cases carried homozygous exon deletions in the *PRKN* gene, and one case carried a heterozygous *GBA1* variant (Table S9). Visual inspection confirmed the presence of these deletions (Supplementary Material).

Outside of the 67 PD-genes, one case also carried a heterozygous deletion in *CACNA1A*, a dystonia gene, removing exons 1 and 2 in a male with LOPD (Table S10). Variants in *CACNA1A* have previously been associated with several neurological disorders, including episodic ataxia type 2, spinocerebellar ataxia type 6, and familial hemiplegic migraine type 1,^62,63^ pointing to its potential relevance for dystonia. The full list of identified SVs is in Table S11.

### PRS for PD are higher in cases

PD cases have significantly higher PRS (Table S12) compared to both 32 internal controls and 1,163 external controls from the GenomeAsia dataset (*P* < 1.44e-34). Our internal set of controls, drawn from healthy individuals from the same host site, show significantly non-zero PRS, pointing to higher background risks for PD from clinical settings.

Consistent with prior studies^64^, individuals with P/LP variants had qualitatively lower PRS relative to those without a genetic diagnosis, though this dataset is not adequately powered to identify significant differences. Cases carrying pathogenic variants are more likely to be driven by rare, high impact mutations rather than a high burden of common risk variants.

### No gene with higher burden of functional variants in cases

PD cases carried significantly more P/LP variants (taken across the union of all genes) compared to controls (Fisher’s test *P* = 0.01), indicating that they carry a higher mutation load. To identify any potential genes enriched in burden among cases, we tested for rare variant gene-based burden in PD cases using gnomAD samples as controls. We used the CoCoRV framework, which incorporates systematic filtering, variant categorization, and ethnicity-stratified association testing while adjusting for multiple comparisons. Although a few genes showed nominal significance (Table S13), none remained significant after false discovery rate (FDR) correction (Figure S5). These findings suggest that the current cohort lacks sufficient statistical power to detect gene-level associations, underscoring the need for larger sample sizes to identify novel candidate genes in PD progression.

## Discussion

In this whole genome-based study of genetic architecture of PD from India, we report a diagnostic yield of 15.73%, with a higher yield in EOPD (27.63%) than LOPD (2.19%). A monogenic cause could be attributed in 50% of familial cases compared to 12.36% of apparently sporadic cases. Pathogenic and likely pathogenic variants were identified in *GBA1 (*45.16% of all identified pathogenic variants)*, PARK7* (9.67%)*, PINK1* (6.45%)*, PLA2G6* (6.45%)*, PRKN* (25.80%)*, SNCA* (3.22%), and *SYNJ1* (3.22%).

*GBA1* variants are important genetic risk factors for PD^61^. Across different populations, the frequency of *GBA1* variants in PD ranges from 3% to 20%^10,65,66^. Rare variants in *GBA1* are present in 5–15% of the Indian population^24^, but many remain unconfirmed as causative for PD^67^. While this may reflect varying importance of the *GBA1* gene in different Indian subpopulations, the choice of reference genome used in analysis may also contribute to under-reporting of *GBA1* variants^68^. In our cohort, *GBA1* alterations were detected in 7.10% of patients. Only a single individual reported a second-degree relative with PD, and all other cases appeared sporadic. As with previous observations^24,69,70^, heterozygous pathogenic *GBA1* variants in our cohort were associated with an earlier age of onset, with 13 of 14 *GBA1* cases presenting with EOPD. The relatively high frequency of these variants in our sample is clinically relevant, as they have been linked to more rapid progression of motor dysfunction and greater cognitive decline^65^.

Three previously undescribed variants in the *PARK7* (*DJ1*) gene were discovered in this study. Two are homozygous splice-site changes identified in individuals with familial PD. The third is a homozygous frameshift variant predicted to be likely pathogenic. In the global literature, *PARK7* mutations are rare, with limited clinical data available on their contribution to early-onset forms of PD^71–73^. Biallelic pathogenic variants in *DJ1* are estimated to account for around 1% of EOPD, frequently associated with diverse or atypical clinical presentations. Consistent with previous reports, all cases identified in our cohort had EOPD, with symptom onset occurring in their late teens to late 30s.

One case with EOPD (onset in the early 20s) had a novel heterozygous missense variant in the *SNCA* gene. *SNCA* mutations are rare in Indian populations and heterozygous missense changes are an uncommon cause of parkinsonism^74,75^. Importantly, only four missense *SNCA* variants are considered definitively pathogenic by databases such as MDSGene. The *SNCA* variant identified in this cohort is classified as “likely pathogenic” by ACMG guidelines, but further functional investigation is warranted to clarify its biological and clinical implications.

Pathogenic biallelic variants in the *PRKN* gene contribute to approximately 10%-20% of EOPD cases globally^76^, although a considerable number of patients are found to carry VUS. Within our study population, eight cases had *PRKN* variants. Of these, one carried a previously undescribed homozygous frameshift mutation, while the other seven had SVs. Seven affected individuals had EOPD, with disease onset ranging from 20 to 30 years, whereas one patient had LOPD. Prior studies indicate ∼62% of *PARK-PRKN* (PARK2) cases manifest as EOPD, with ∼16% presenting as juvenile-onset PD^77^. Exon 3 is affected in five out of seven patients with *PRKN* exon deletions. Exons 3-8 of *PRKN* span the central core of FRA6E, a well-established mutation hotspot and the likely mechanistic basis for the clustering of SVs in this region^78^.

Two EOPD patients carry known biallelic pathogenic variants in *PINK1*, one with a missense mutation and the other with a frameshift mutation. Two additional cases carry compound heterozygous variants in *PLA2G6*; one case carries (*p.Ala781Thr* and *p.Arg747Trp*), the latter being a known pathogenic variant supported by functional studies. Another case carried (p.Arg741Gln and p.Ala633Val), both of which are pathogenic. Another PD case carried a known homozygous missense variant in *SYNJ1*, with symptom onset at early 30s. We also identified two cases with a novel pathogenic variant in *ATXN2*. Although *ATXN2* is not classified as a PD gene, multiple studies have shown that CAG-repeat expansions (particularly 33–43 repeats) can have levodopa-responsive parkinsonism in addition to the full SCA2 spectrum^79^. Large cohort studies and meta-analyses further highlight ethnic variability and pleiotropic effects, linking *ATXN2* to PD, ALS, and SCA2^80–82^. This suggests that *ATXN2* may play a broader role in neurodegeneration, with potential relevance for parkinsonism.

We also identified many VUS in PD genes; among them one *LRRK2* variant (p.Ala1442Pro) was found to be highly damaging and predicted to be pathogenic by AlphaMissense. This variant has previously been reported in an Australian familial PD case as a novel mutation associated with levodopa-responsive parkinsonism,^83^ and more recently, functional studies demonstrated that p.Ala1442Pro increases *LRRK2* kinase activity and alters microtubule binding^84^, supporting its pathogenic potential. Globally, the genes with VUS in PD cases are enriched for neurodegeneration pathways; this strongly suggests that the VUS set includes many likely causal variants that require more functional evidence and better variant prioritization. The identification of rare variants in our cohort underscores the need for comprehensive genetic analysis and careful phenotyping to improve our understanding of genotype-phenotype relationships and enhance early diagnosis, risk assessment, and tailored management strategies for PD patients in diverse populations.

Our results demonstrate that European-derived PRS maintain predictive value in Indian populations. This suggests that the overall polygenic contribution to PD risk is conserved across populations despite differences in individual risk variants and allele frequencies. Controls drawn from the same clinical center (one focused on mental health and neuroscience) show significantly higher PRS than external controls, perhaps pointing to the shared genetic architecture between neurological, psychiatric, and neurodegenerative phenotypes. This also points to potentially higher PD risk for carriers of other brain-related disorders^85^.

The Indian population remains under-represented in PD genetic research, despite the high burden of the disease. Compared to European cohorts, where population homogeneity and easier patient recruitment have facilitated large-scale studies, insights into PD genetics in India are still limited. Given that populations differ in allele frequencies, linkage disequilibrium patterns, and effect estimates, no single population can fully elucidate the genetic underpinnings of PD. This underscores the scientific and clinical imperative to expand genetic research in diverse populations such as India, to capture population-specific risk variants and enhance understanding of the global genetic architecture of PD. Genes with likely pathogenic variants in cases are enriched in lysosomal pathways, underscoring the relevance of PD etiology^86^.

Finally, while our current study is underpowered to detect any genome-wide significant associations, this cohort provides a valuable foundation for larger collaborative studies that could identify novel PD loci specific to Indian populations. We also demonstrate the clinical utility of comprehensive genetic screening in Indian PD patients and highlight the need for population-specific diagnostic approaches.

## Acknowledgements

We acknowledge funding from the SKAN research trust. We thank all participants in the study. We thank Mohammad Hanif Kaba Mujawar and Vinayak Hosawad for generating high-quality sequencing data, and Nidhi Gambhir for supporting the SV analysis. We also acknowledge Anand Kumar and Jothibasu V for IT support and CBR’s HPC facility for computational resources.

## Data Availability

All the raw sequence files for the 31 diagnosed cases will be available to dbGaP upon publication.

## Author contributions

Design: A.S, L.D, P.K.P

Execution: All authors

Analysis: H.A, A.P, C.S, B.K.P, A.S, S.R

Writing: All authors

Editing of the final version of the manuscript: H.A, A.S, L.D, S.R

## Funding agencies

The study was supported by the SKAN research trust.

---

**Figure S1.**
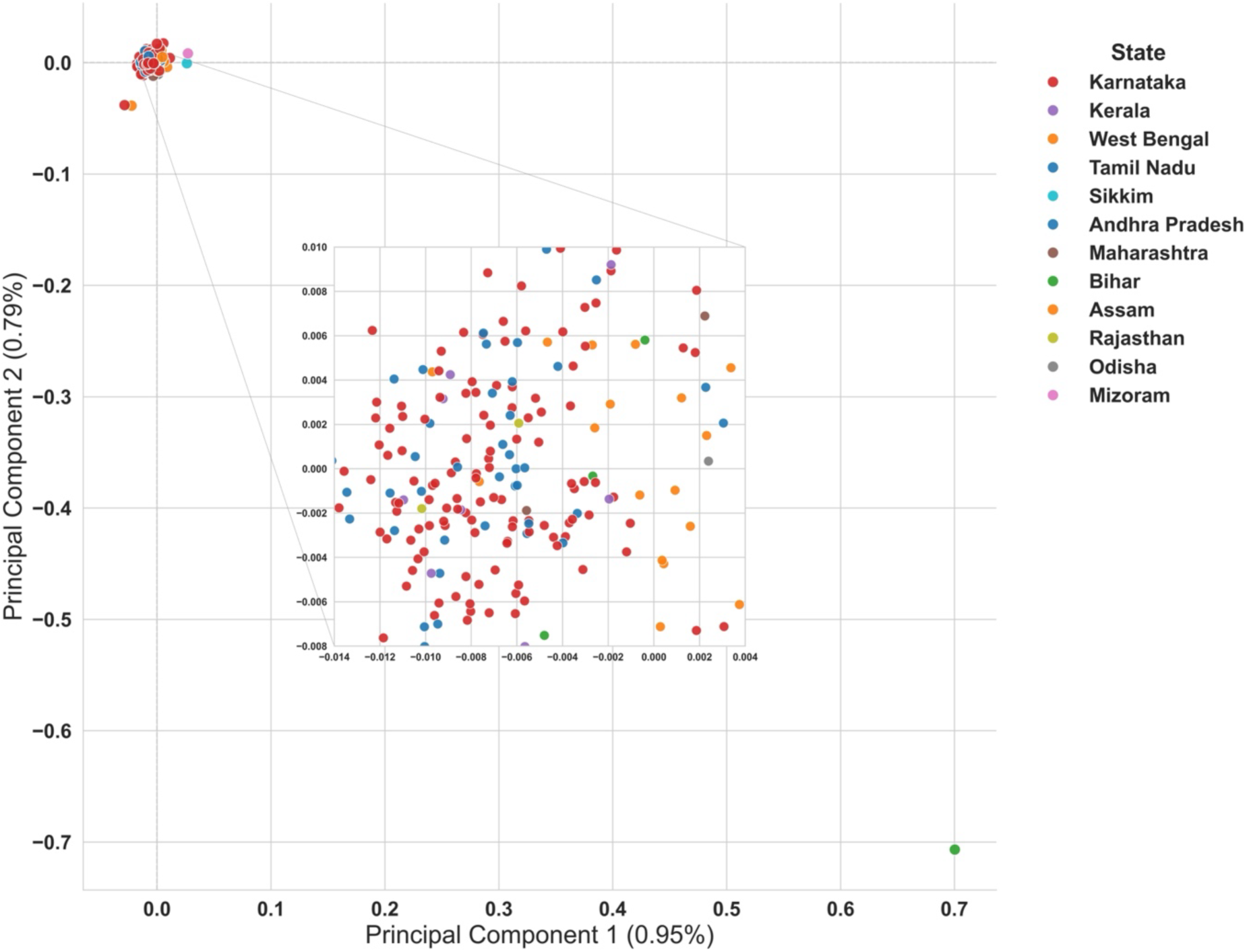
PCA plot of SNP data from SKAN cohort, colored by state. Principal component analysis (PCA) of SNP genotype data from all unrelated individuals. The plot shows PC1 vs PC2, which explain 0.95% and 0.79% of the total genetic variance, respectively. Each point represents an individual, colored by their state. The clustering along PC1 or PC2 reflects major sources of genetic structure in the dataset, potentially due to population stratification, geographic origin, or phenotypic differentiation.

**Figure S2.**
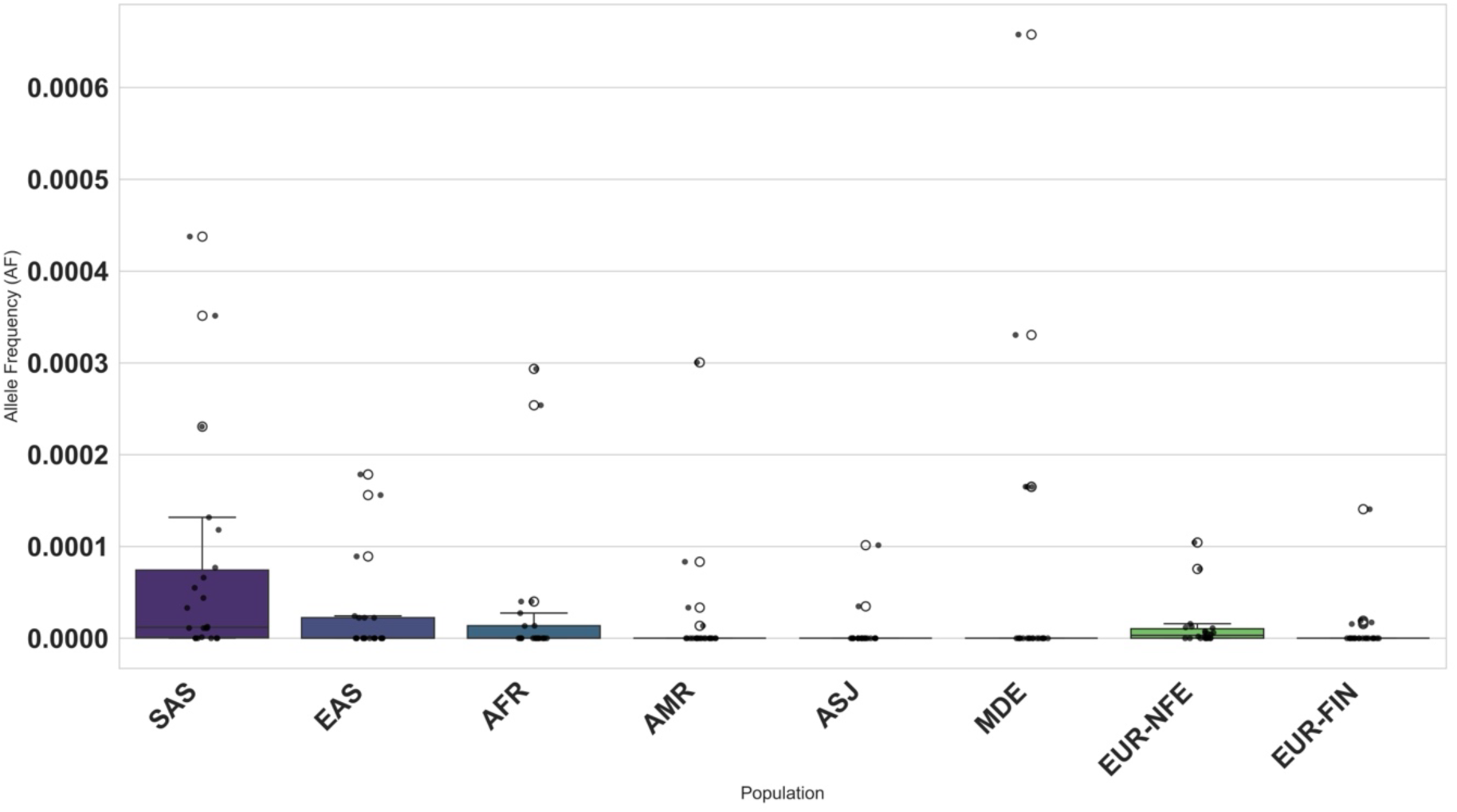
Mean Allele Frequencies of Pathogenic Variants in PD Genes Across Populations. Bar plot showing the mean allele frequency (AF) of pathogenic variants identified in known Parkinson’s disease (PD) genes, stratified by population. The Y-axis indicates the mean AF, and the X-axis shows the population groups. Each bar represents the average frequency of pathogenic alleles within that group. This comparison highlights potential population-level differences in the burden of disease-associated variants.

**Figure S3.**
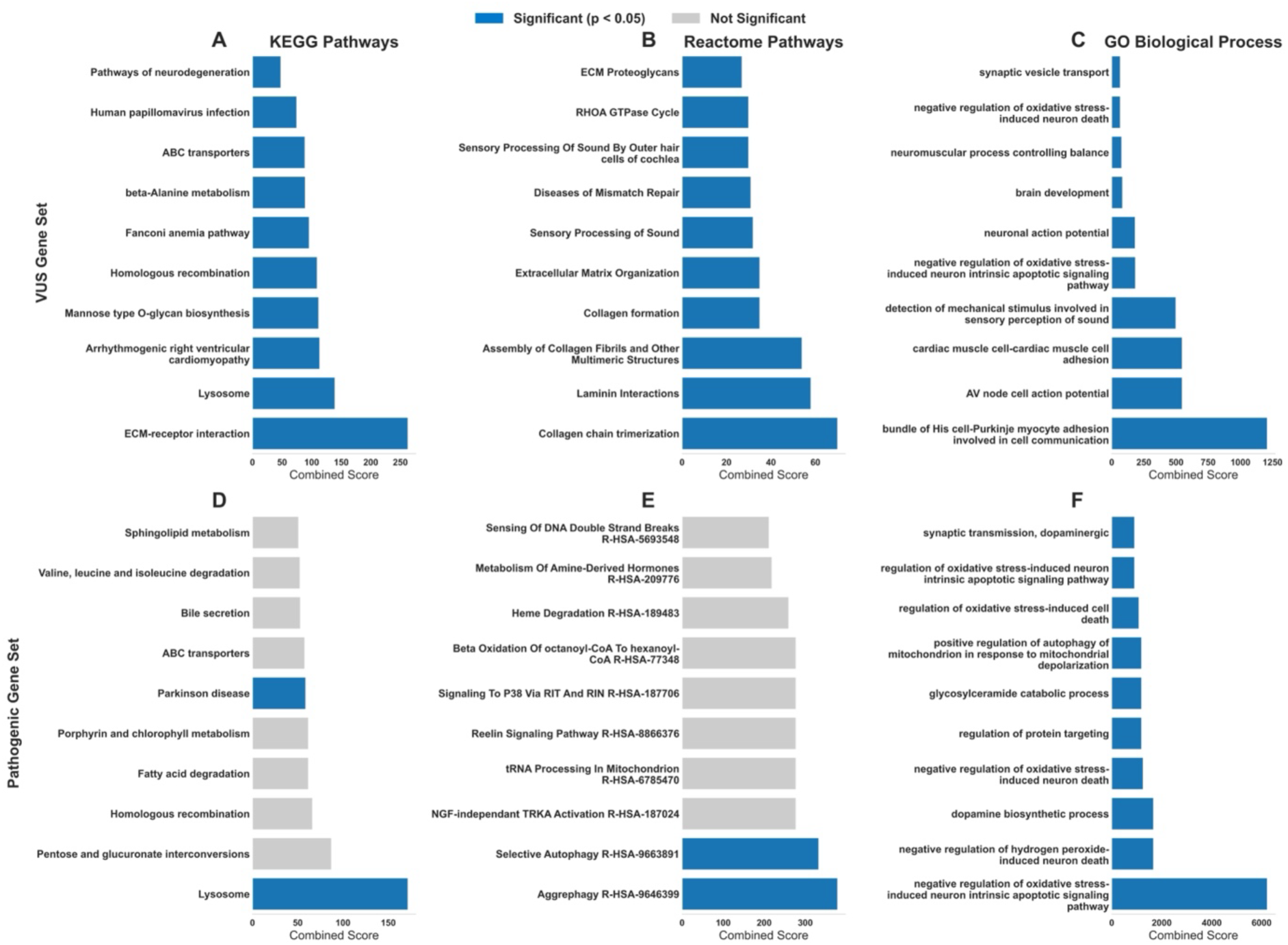
Pathway enrichment analysis of genes with variants of uncertain significance (VUS) and likely pathogenic/pathogenic variants. Top enriched pathways for the VUS gene set (top row, A-C) and the pathogenic gene set (bottom row, D-F). Analyses were performed using three databases: KEGG (A, D), Reactome (B, E), and GO Biological Process (C, F).

**Figure S4.**
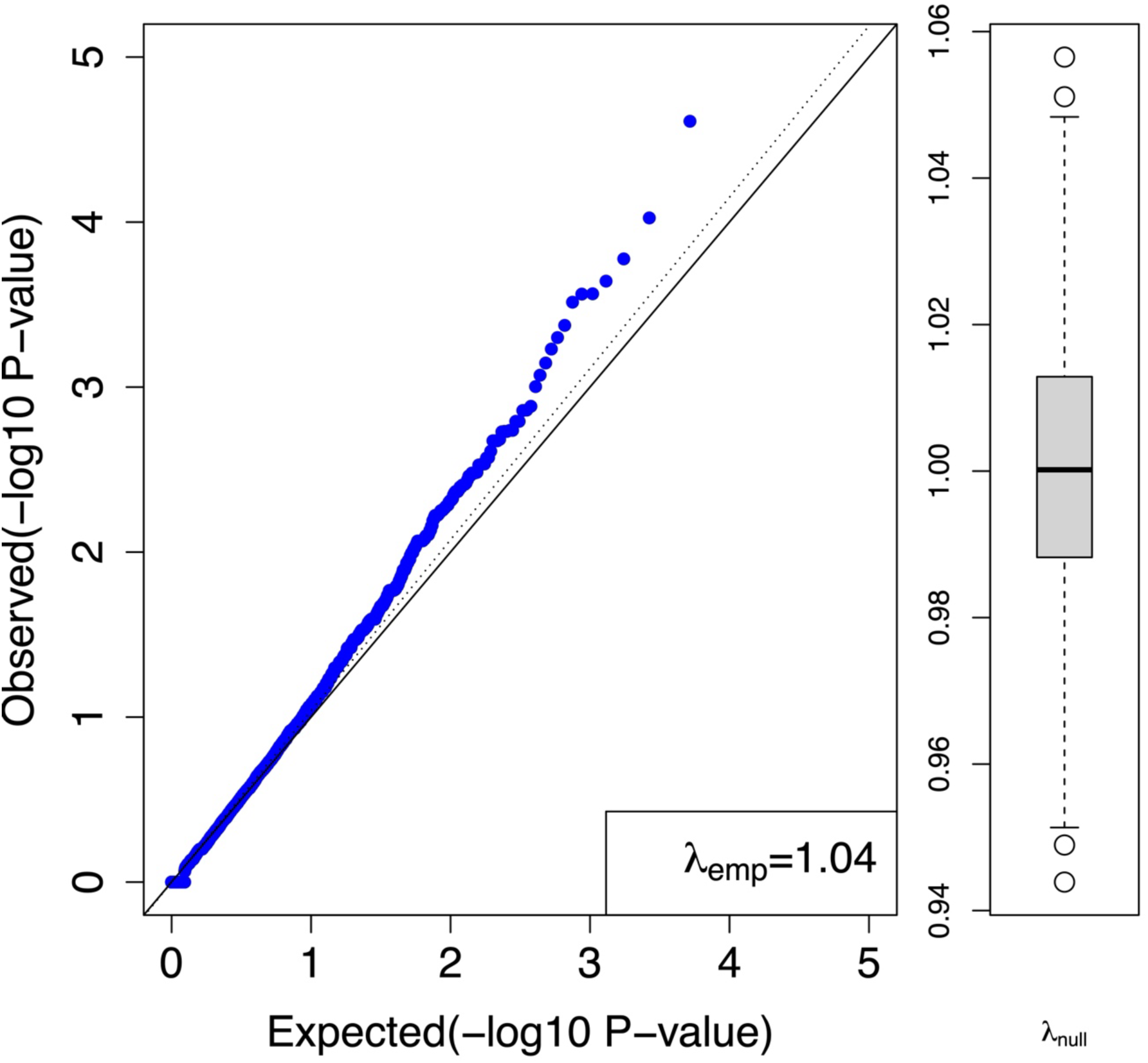
Quantile-Quantile (QQ) Plot of Gene-Based Burden Testing in PD. QQ plot illustrating the distribution of observed versus expected p-values from gene-based rare variant burden testing using the CoCoRV framework (λ = 1.04). The slight deviation from the null suggests potential signals, but the modest inflation factor and limited number of significant hits indicate that the analysis remains underpowered due to the small sample size (n = 197 PD cases).

## Table Legends

**Table S1.** Cohort Description

**Table S2.** List of 67 genes previously associated with PD, and 37 genes associated with dystonia and parkinsonism

**Table S3.** List of pathogenic variants in PD associated genes

**Table S4.** List of VUS in PD genes

**Table S5.** List of pathogenic variants in parkinsonism and dystonia genes

**Table S6.** List of all variants in parkinsonism and dystonia genes

**Table S7.** List of VUS in non-PD genes

**Table S8.** List of pathogenic mutations in non-PD genes

**Table S9.** List of SVs in PD associated genes

**Table S10.** List of SVs in parkinsonism and dystonia genes

**Table S11.** List of SVs in non-PD genes

**Table S12.** PRS values

**Table S13.** Gene burden analysis results

**Table S14.** Pathway enrichment analysis results

## References

1. Bloem BR, Okun MS, Klein C. Parkinson’s disease. The Lancet. 2021 June 12;397(10291):2284–303.

2. Ben-Shlomo Y, Darweesh S, Llibre-Guerra J, Marras C, Luciano MS, Tanner C. The epidemiology of Parkinson’s disease. Lancet. 2024 Jan 20;403(10423):283–92.

3. Dorsey ER, Bloem BR. The Parkinson Pandemic-A Call to Action. JAMA Neurol. 2018 Jan 1;75(1):9–10.

4. Dorsey ER, Sherer T, Okun MS, Bloem BR. The Emerging Evidence of the Parkinson Pandemic. J Parkinsons Dis. 8(Suppl 1):S3–8.

5. Blauwendraat C, Nalls MA, Singleton AB. The genetic architecture of Parkinson’s disease. The Lancet Neurology. 2020 Feb 1;19(2):170–8.

6. Deng H, Wang P, Jankovic J. The genetics of Parkinson disease. Ageing Research Reviews. 2018 Mar 1;42:72–85.

7. Deng H, Wang P, Jankovic J. The genetics of Parkinson disease. Ageing Research Reviews. 2018 Mar 1;42:72–85.

8. Pitz V, Makarious MB, Bandres-Ciga S, Iwaki H, Singleton AB, Nalls M, et al. Analysis of rare Parkinson’s disease variants in millions of people. npj Parkinsons Dis. 2024 Jan 8;10(1):11.

9. Towns C, Fang ZH, Tan MMX, Jasaityte S, Schmaderer TM, Stafford EJ, et al. Parkinson’s families project: a UK-wide study of early onset and familial Parkinson’s disease. npj Parkinsons Dis. 2024 Oct 17;10(1):188.

10. Bhowmick SS, Desai SD. Genetic Sketch of Parkinson’s Disease in India. Ann Indian Acad Neurol. 2025;28(4):495–504.

11. Nalls MA, Blauwendraat C, Vallerga CL, Heilbron K, Bandres-Ciga S, Chang D, et al. Identification of novel risk loci, causal insights, and heritable risk for Parkinson’s disease: a meta-analysis of genome-wide association studies. Lancet Neurol. 2019 Dec;18(12):1091–102.

12. Nishioka K, Imai Y, Yoshino H, Li Y, Funayama M, Hattori N. Clinical Manifestations and Molecular Backgrounds of Parkinson’s Disease Regarding Genes Identified From Familial and Population Studies. Front Neurol. 2022;13:764917.

13. Nishioka K, Imai Y, Yoshino H, Li Y, Funayama M, Hattori N. Clinical Manifestations and Molecular Backgrounds of Parkinson’s Disease Regarding Genes Identified From Familial and Population Studies. Front Neurol. 2022;13:764917.

14. Nalls MA, Blauwendraat C, Vallerga CL, Heilbron K, Bandres-Ciga S, Chang D, et al. Identification of novel risk loci, causal insights, and heritable risk for Parkinson’s disease: a meta-analysis of genome-wide association studies. The Lancet Neurology. 2019 Dec 1;18(12):1091–102.

15. Kline EM, Houser MC, Herrick MK, Seibler P, Klein C, West A, et al. Genetic and Environmental Factors in Parkinson’s Disease Converge on Immune Function and Inflammation. Mov Disord. 2021 Jan;36(1):25–36.

16. Kolicheski A, Turcano P, Tamvaka N, McLean PJ, Springer W, Savica R, et al. Early-Onset Parkinson’s Disease: Creating the Right Environment for a Genetic Disorder. J Parkinsons Dis. 12(8):2353–67.

17. Polito L, Greco A, Seripa D. Genetic Profile, Environmental Exposure, and Their Interaction in Parkinson’s Disease. Parkinsons Dis. 2016;2016:6465793.

18. Tsalenchuk M, Gentleman SM, Marzi SJ. Linking environmental risk factors with epigenetic mechanisms in Parkinson’s disease. npj Parkinsons Dis. 2023 Aug 25;9(1):123.

19. Rajan R, Divya KP, Kandadai RM, Yadav R, Satagopam VP, Madhusoodanan UK, et al. Genetic Architecture of Parkinson’s Disease in the Indian Population: Harnessing Genetic Diversity to Address Critical Gaps in Parkinson’s Disease Research. Front Neurol. 2020 June 18;11:524.

20. Kim JJ, Vitale D, Otani DV, Lian MM, Heilbron K, Iwaki H, et al. Multi-ancestry genome-wide association meta-analysis of Parkinson’s disease. Nat Genet. 2024 Jan;56(1):27–36.

21. Koros C, Bougea A, Simitsi AM, Papagiannakis N, Angelopoulou E, Pachi I, et al. The Landscape of Monogenic Parkinson’s Disease in Populations of Non-European Ancestry: A Narrative Review. Genes (Basel). 2023 Nov 17;14(11):2097.

22. Nishioka K, Imai Y, Yoshino H, Li Y, Funayama M, Hattori N. Clinical Manifestations and Molecular Backgrounds of Parkinson’s Disease Regarding Genes Identified From Familial and Population Studies. Front Neurol. 2022 June 2;13:764917.

23. Dehestani M, Liu H, Gasser T. Polygenic Risk Scores Contribute to Personalized Medicine of Parkinson’s Disease. J Pers Med. 2021 Oct 15;11(10):1030.

24. Kamath SD, Phulpagar P, Holla VV, Kamble N, Yadav R, Muthusamy B, et al. Genetic architecture of a single cohort of 230 Indian Parkinson’s Disease patients. Parkinsonism & Related Disorders. 2024 Dec;129:107157.

25. Kumar S, Yadav N, Pandey S, Muthane UB, Govindappa ST, Abbas MM, et al. Novel and reported variants in Parkinson’s disease genes confer high disease burden among Indians. Parkinsonism Relat Disord. 2020 Sept;78:46–52.

26. Soldner F, Stelzer Y, Shivalila CS, Abraham BJ, Latourelle JC, Barrasa MI, et al. Parkinson-associated risk variant in distal enhancer of α-synuclein modulates target gene expression. Nature. 2016 May 5;533(7601):95–9.

27. Boussaad I, Obermaier CD, Hanss Z, Bobbili DR, Bolognin S, Glaab E, et al. A patient-based model of RNA mis-splicing uncovers treatment targets in Parkinson’s disease. Science Translational Medicine. 2020 Sept 9;12(560):eaau3960.

28. Cho N, Joo J, Choi S, Kang BG, Lee AJ, Youn SY, et al. A novel splicing variant of DJ-1 in Parkinson’s disease induces mitochondrial dysfunction. Heliyon. 2023 Mar 1;9(3):e14039.

29. Lu Y, Li M, Gao Z, Ma H, Chong Y, Hong J, et al. Advances in Whole Genome Sequencing: Methods, Tools, and Applications in Population Genomics. Int J Mol Sci. 2025 Jan 4;26(1):372.

30. Liu X, Hu F, Zhang D, Li Z, He J, Zhang S, et al. Whole genome sequencing enables new genetic diagnosis for inherited retinal diseases by identifying pathogenic variants. NPJ Genom Med. 2024 Jan 20;9:6.

31. Postuma RB, Berg D, Stern M, Poewe W, Olanow CW, Oertel W, et al. MDS clinical diagnostic criteria for Parkinson’s disease. Movement Disorders. 2015;30(12):1591– 601.

32. FastQC: A quality control tool for high throughput sequence data – ScienceOpen [Internet]. [cited 2025 Oct 16]. Available from: https://www.scienceopen.com/document?vid=de674375-ab83-4595-afa9-4c8aa9e4e736

33. Ewels P, Magnusson M, Lundin S, Käller M. MultiQC: summarize analysis results for multiple tools and samples in a single report. Bioinformatics. 2016 Oct 1;32(19):3047–8.

34. Bolger AM, Lohse M, Usadel B. Trimmomatic: a flexible trimmer for Illumina sequence data. Bioinformatics. 2014 Aug 1;30(15):2114–20.

35. Vasimuddin Md, Misra S, Li H, Aluru S. Efficient Architecture-Aware Acceleration of BWA-MEM for Multicore Systems. In: 2019 IEEE International Parallel and Distributed Processing Symposium (IPDPS) [Internet]. 2019 [cited 2025 Oct 16]. p. 314–24. Available from: https://ieeexplore.ieee.org/document/8820962

36. Poplin R, Ruano-Rubio V, Depristo MA, Fennell TJ, Carneiro MO, Geraldine A Van Der Auwera, et al. Scaling accurate genetic variant discovery to tens of thousands of samples. bioRxiv. 1.3. 2018;

37. Cingolani P, Platts A, Wang LL, Coon M, Nguyen T, Wang L, et al. A program for annotating and predicting the effects of single nucleotide polymorphisms, SnpEff. Fly (Austin). 2012 Apr 1;6(2):80–92.

38. Wang K, Li M, Hakonarson H. ANNOVAR: functional annotation of genetic variants from high-throughput sequencing data. Nucleic Acids Res. 2010 Sept 1;38(16):e164.

39. Karczewski KJ, Francioli LC, Tiao G, Cummings BB, Alföldi J, Wang Q, et al. The mutational constraint spectrum quantified from variation in 141,456 humans. Nature. 2020 May;581(7809):434–43.

40. Jaganathan K, Kyriazopoulou Panagiotopoulou S, McRae JF, Darbandi SF, Knowles D, Li YI, et al. Predicting Splicing from Primary Sequence with Deep Learning. Cell. 2019 Jan 24;176(3):535–548.e24.

41. Richards S, Aziz N, Bale S, Bick D, Das S, Gastier-Foster J, et al. Standards and guidelines for the interpretation of sequence variants: a joint consensus recommendation of the American College of Medical Genetics and Genomics and the Association for Molecular Pathology. Genet Med. 2015 May;17(5):405–23.

42. Abyzov A, Urban AE, Snyder M, Gerstein M. CNVnator: An approach to discover, genotype, and characterize typical and atypical CNVs from family and population genome sequencing. Genome Res. 2011 June;21(6):974–84.

43. Layer RM, Chiang C, Quinlan AR, Hall IM. LUMPY: a probabilistic framework for structural variant discovery. Genome Biology. 2014 June 26;15(6):R84.

44. Chen X, Schulz-Trieglaff O, Shaw R, Barnes B, Schlesinger F, Källberg M, et al. Manta: rapid detection of structural variants and indels for germline and cancer sequencing applications. Bioinformatics. 2016 Apr 15;32(8):1220–2.

45. Rausch T, Zichner T, Schlattl A, Stütz AM, Benes V, Korbel JO. DELLY: structural variant discovery by integrated paired-end and split-read analysis. Bioinformatics. 2012 Sept 15;28(18):i333–9.

46. Fan X, Abbott TE, Larson D, Chen K. BreakDancer – Identification of Genomic Structural Variation from Paired-End Read Mapping. Curr Protoc Bioinformatics. 2014;2014:10.1002/0471250953.bi1506s45.

47. Rajaby R, Liu DX, Au CH, Cheung YT, Lau AYT, Yang QY, et al. INSurVeyor: improving insertion calling from short read sequencing data. Nat Commun. 2023 June 5;14(1):3243.

48. Jeffares DC, Jolly C, Hoti M, Speed D, Shaw L, Rallis C, et al. Transient structural variations have strong effects on quantitative traits and reproductive isolation in fission yeast. Nat Commun. 2017 Jan 24;8(1):14061.

49. Robinson JT, Thorvaldsdóttir H, Winckler W, Guttman M, Lander ES, Getz G, et al. Integrative genomics viewer. Nat Biotechnol. 2011 Jan;29(1):24–6.

50. Privé F, Arbel J, Vilhjálmsson BJ. LDpred2: better, faster, stronger. Bioinformatics. 2021 Apr 1;36(22–23):5424–31.

51. Choi SW, Mak TSH, O’Reilly PF. Tutorial: a guide to performing polygenic risk score analyses. Nat Protoc. 2020 Sept;15(9):2759–72.

52. Privé F, Albiñana C, Arbel J, Pasaniuc B, Vilhjálmsson BJ. Inferring disease architecture and predictive ability with LDpred2-auto. The American Journal of Human Genetics. 2023 Dec 7;110(12):2042–55.

53. Wall JD, Stawiski EW, Ratan A, Kim HL, Kim C, Gupta R, et al. The GenomeAsia 100K Project enables genetic discoveries across Asia. Nature. 2019 Dec;576(7785):106–11.

54. Saleheen D, Natarajan P, Armean IM, Zhao W, Rasheed A, Khetarpal SA, et al. Human knockouts and phenotypic analysis in a cohort with a high rate of consanguinity. Nature. 2017 Apr;544(7649):235–9.

55. Chen EY, Tan CM, Kou Y, Duan Q, Wang Z, Meirelles GV, et al. Enrichr: interactive and collaborative HTML5 gene list enrichment analysis tool. BMC Bioinformatics. 2013 Apr 15;14:128.

56. Chen W, Wang S, Tithi SS, Ellison DW, Schaid DJ, Wu G. A rare variant analysis framework using public genotype summary counts to prioritize disease-predisposition genes. Nat Commun. 2022 May 11;13(1):2592.

57. Girija MS, Kishore A. Clinical approach to monogenic Parkinson’s disease. Annals of Movement Disorders. 2025 Apr;8(1):1.

58. Bonato G, Antonini A, Pistonesi F, Campagnolo M, Guerra A, Biundo R, et al. Genetic mutations in Parkinson’s disease: screening of a selected population from North-Eastern Italy. Neurol Sci. 2025;46(1):165–74.

59. Rajan R, Holla VV, Kamble N, Yadav R, Pal PK. Genetic heterogeneity of early onset Parkinson disease: The dilemma of clinico-genetic correlation. Parkinsonism & Related Disorders. 2024 Dec 1;129:107146.

60. Yadav R, Kapoor S, Madhukar M, Naduthota RM, Kumar A, Pal PK. Genetic analysis of the glucocerebrosidase gene in South Indian patients with Parkinson’s disease. Neurol India. 2018;66(6):1649–54.

61. Cheng J, Novati G, Pan J, Bycroft C, Žemgulytė A, Applebaum T, et al. Accurate proteome-wide missense variant effect prediction with AlphaMissense. Science. 2023 Sept 22;381(6664):eadg7492.

62. Isaacs DA, Bradshaw MJ, Brown K, Hedera P. Case report of novel CACNA1A gene mutation causing episodic ataxia type 2. SAGE Open Med Case Rep. 2017 May 8;5:2050313X17706044.

63. Lipman AR, Fan X, Shen Y, Chung WK. Clinical and genetic characterization of CACNA1A-related disease. Clin Genet. 2022 Oct;102(4):288–95.

64. Goldstein O, Shani S, Gana-Weisz M, Elkoshi N, Casey F, Sun YH, et al. The effect of polygenic risk score on PD risk and phenotype in LRRK2 G2019S and GBA1 carriers. Journal of Parkinson’s Disease. 2025 Mar 1;15(2):291–9.

65. Sidransky E, Nalls MA, Aasly JO, Aharon-Peretz J, Annesi G, Barbosa ER, et al. Multicenter analysis of glucocerebrosidase mutations in Parkinson’s disease. N Engl J Med. 2009 Oct 22;361(17):1651–61.

66. Alcalay RN, Dinur T, Quinn T, Sakanaka K, Levy O, Waters C, et al. Comparison of Parkinson risk in Ashkenazi Jewish patients with Gaucher disease and GBA heterozygotes. JAMA Neurol. 2014 June;71(6):752–7.

67. Andrews SV, Kukkle PL, Menon R, Geetha TS, Goyal V, Kandadai RM, et al. The Genetic Drivers of Juvenile, Young, and Early-Onset Parkinson’s Disease in India. Mov Disord. 2024 Feb;39(2):339–49.

68. Jia T, Munson B, Lango Allen H, Ideker T, Majithia AR. Thousands of missing variants in the UK Biobank are recoverable by genome realignment. Annals of Human Genetics. 2020 May;84(3):214–20.

69. Granek Z, Barczuk J, Siwecka N, Rozpędek-Kamińska W, Kucharska E, Majsterek I. GBA1 Gene Mutations in α-Synucleinopathies—Molecular Mechanisms Underlying Pathology and Their Clinical Significance. International Journal of Molecular Sciences. 2023 Jan;24(3):2044.

70. Koros C, Bougea A, Alefanti I, Simitsi AM, Papagiannakis N, Pachi I, et al. A Global Perspective of GBA1-Related Parkinson’s Disease: A Narrative Review. Genes. 2024 Dec;15(12):1605.

71. Stephenson SE, Djaldetti R, Rafehi H, Wilson GR, Gillies G, Bahlo M, et al. Familial early onset Parkinson’s disease caused by a homozygous frameshift variant in PARK7: Clinical features and literature update. Parkinsonism Relat Disord. 2019 July;64:308–11.

72. Alcalay RN, Caccappolo E, Mejia-Santana H, Tang MX, Rosado L, Ross BM, et al. Frequency of Known Mutations in Early-Onset Parkinson Disease: Implication for Genetic Counseling: The Consortium on Risk for Early Onset Parkinson Disease Study. Arch Neurol. 2010 Sept 1;67(9):1116–22.

73. Koziorowski D, Hoffman-Zacharska D, Sławek J, Jamrozik Z, Janik P, Potulska-Chromik A, et al. Incidence of mutations in the PARK2, PINK1, PARK7 genes in Polish early-onset Parkinson disease patients. Neurol Neurochir Pol. 2013;47(4):319–24.

74. Vishwanathan Padmaja M, Jayaraman M, Srinivasan AV, Srikumari Srisailapathy CR, Ramesh A. The SNCA (A53T, A30P, E46K) and LRRK2 (G2019S) mutations are rare cause of Parkinson’s disease in South Indian patients. Parkinsonism & Related Disorders. 2012 July 1;18(6):801–2.

75. Kishore A, Sturm M, Soman Pillai K, Hakkaart C, Kalikavil Puthanveedu D, Urulangodi M, et al. Resequencing the complete SNCA locus in Indian patients with Parkinson’s disease. npj Parkinsons Dis. 2024 Apr 15;10(1):85.

76. Klein C, Lohmann-Hedrich K. Impact of recent genetic findings in Parkinson’s disease. Curr Opin Neurol. 2007 Aug 1;20(4):453–64.

77. Kolicheski A, Turcano P, Tamvaka N, McLean PJ, Springer W, Savica R, et al. Early-Onset Parkinson’s Disease: Creating the Right Environment for a Genetic Disorder. J Parkinsons Dis. 12(8):2353–67.

78. Denison SR, Callahan G, Becker NA, Phillips LA, Smith DI. Characterization of FRA6E and its potential role in autosomal recessive juvenile parkinsonism and ovarian cancer. Genes Chromosomes Cancer. 2003 Sept;38(1):40–52.

79. Van Damme P, Veldink JH, van Blitterswijk M, Corveleyn A, van Vught PWJ, Thijs V, et al. Expanded ATXN2 CAG repeat size in ALS identifies genetic overlap between ALS and SCA2. Neurology. 2011 June 14;76(24):2066–72.

80. Casse F, Courtin T, Tesson C, Ferrien M, Noël S, Fauret-Amsellem A, et al. Detection of ATXN2 Expansions in an Exome Dataset: An Underdiagnosed Cause of Parkinsonism. Mov Disord Clin Pract. 2023 Mar 7;10(4):664–9.

81. Demaegd KC, Kernan A, Cooper-Knock J, van Vugt JJFA, Harvey C, Moll T, et al. An observational study of pleiotropy and penetrance of amyotrophic lateral sclerosis associated with CAG-repeat expansion of ATXN2. Eur J Hum Genet. 2025 Sept;33(9):1106–12.

82. Costa RG, Conceição A, Matos CA, Nóbrega C. The polyglutamine protein ATXN2: from its molecular functions to its involvement in disease. Cell Death Dis. 2024 June 14;15(6):415.

83. Huang Y, Halliday GM, Vandebona H, Mellick GD, Mastaglia F, Stevens J, et al. Prevalence and clinical features of common LRRK2 mutations in Australians with Parkinson’s disease. Mov Disord. 2007 May 15;22(7):982–9.

84. Kalogeropulou AF, Purlyte E, Tonelli F, Lange SM, Wightman M, Prescott AR, et al. Impact of 100 LRRK2 variants linked to Parkinson’s disease on kinase activity and microtubule binding. Biochem J. 2022 Sept 16;479(17):1759–83.

85. Wingo TS, Liu Y, Gerasimov ES, Vattathil SM, Wynne ME, Liu J, et al. Shared mechanisms across the major psychiatric and neurodegenerative diseases. Nat Commun. 2022 July 26;13(1):4314.

86. Robak LA, Jansen IE, van Rooij J, Uitterlinden AG, Kraaij R, Jankovic J, et al. Excessive burden of lysosomal storage disorder gene variants in Parkinson’s disease. Brain. 2017 Dec;140(12):3191–203.

