## Supplementary material for "Whole-genome sequencing of 197 cases with Parkinson’s Disease reveals novel pathogenic variants in the Indian population"

**Contents:**

1. IGV snapshots of SV deletions

2. Supplementary Table column descriptions

***IGV Snapshots***

1. *PRKN* deletions:

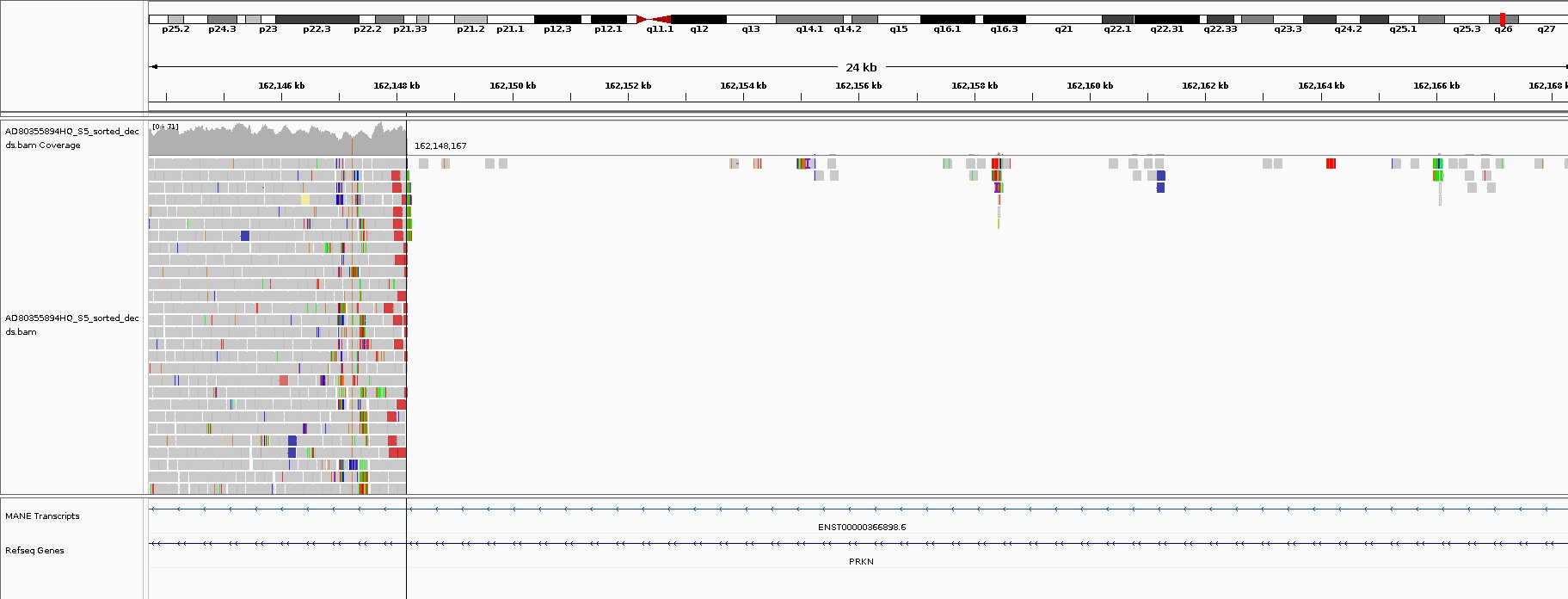

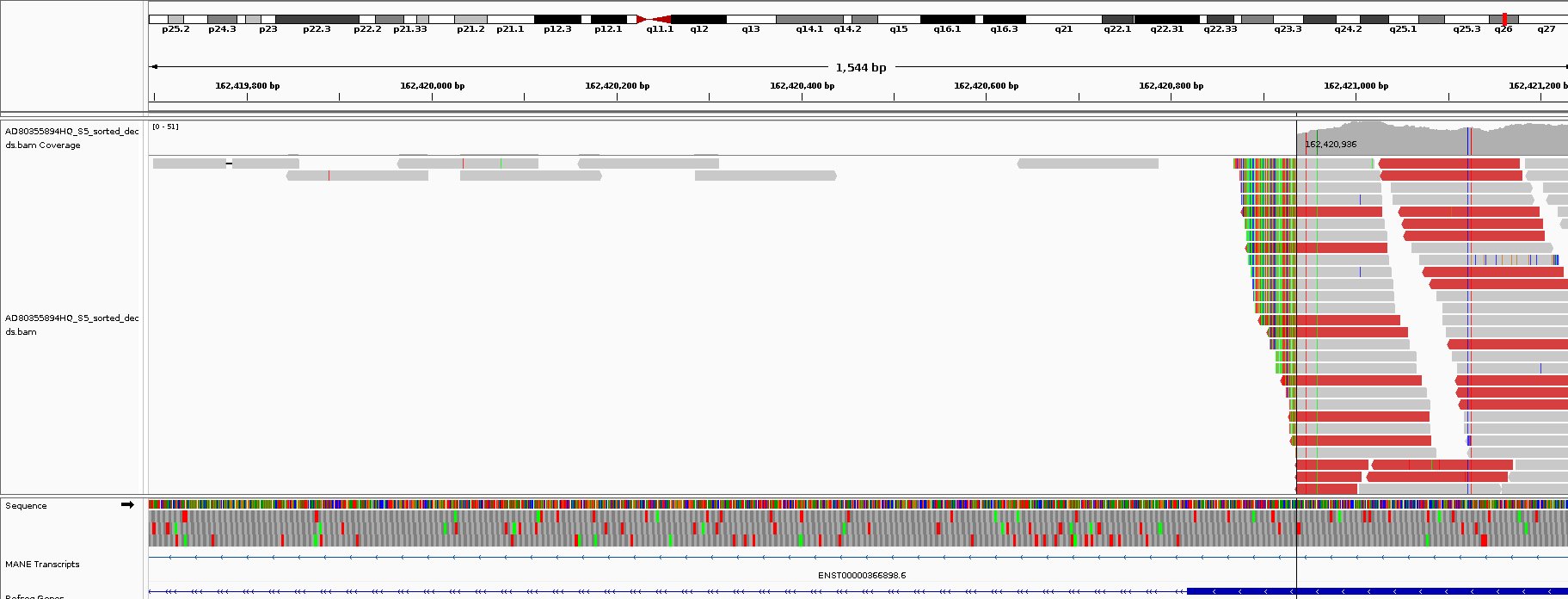

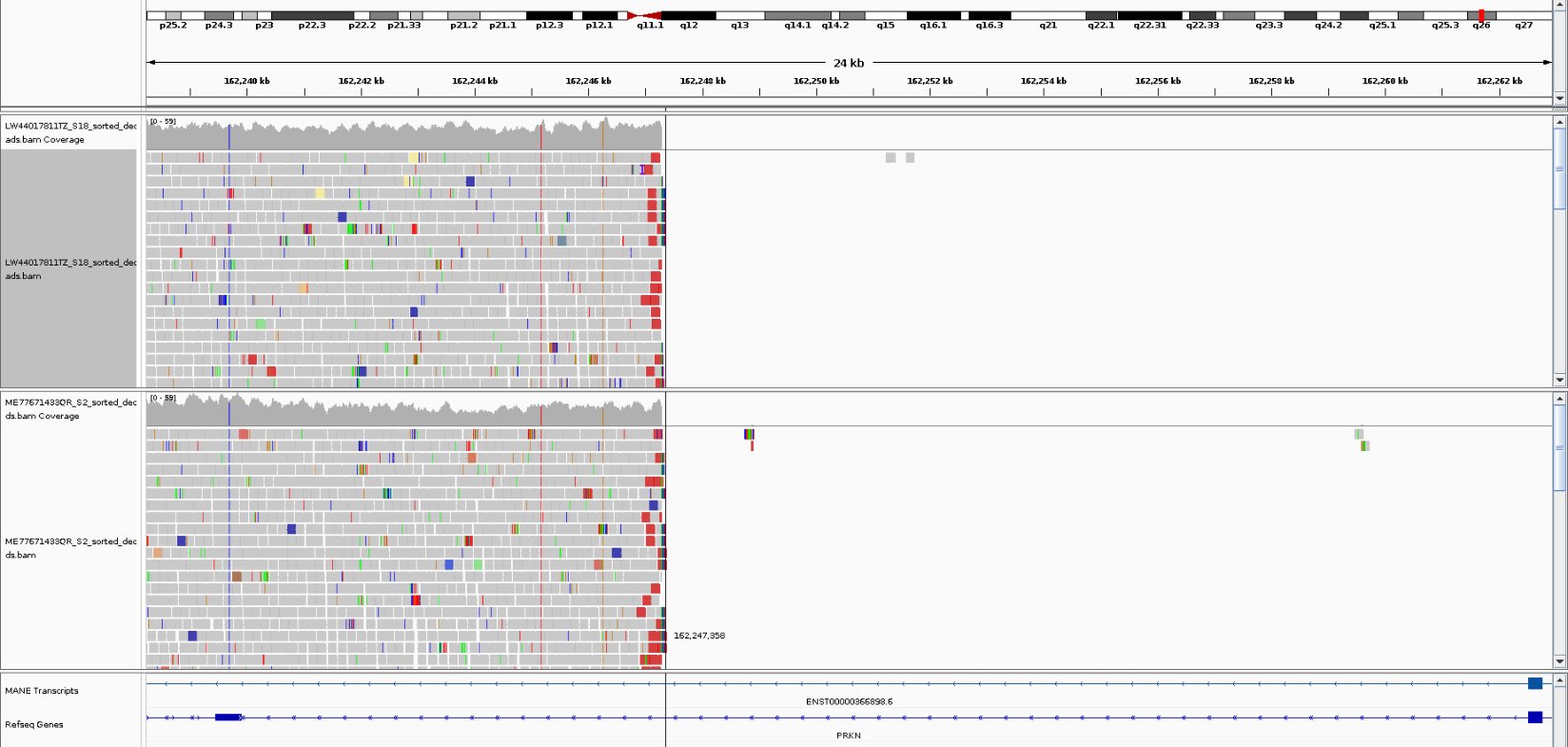

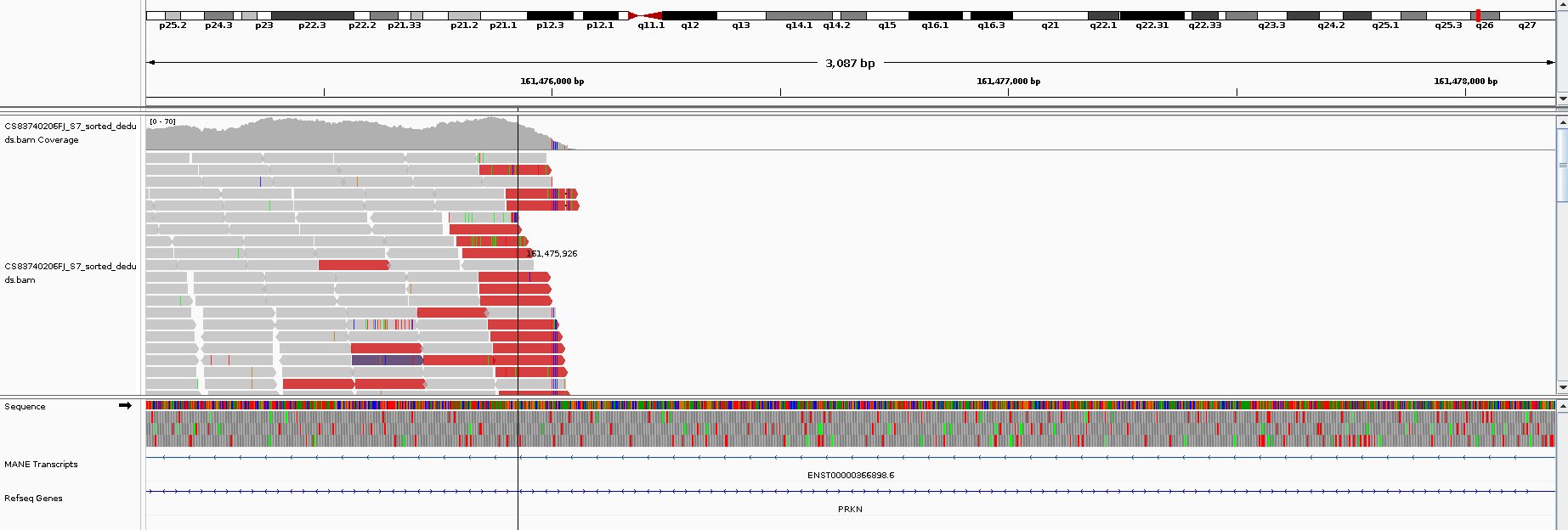

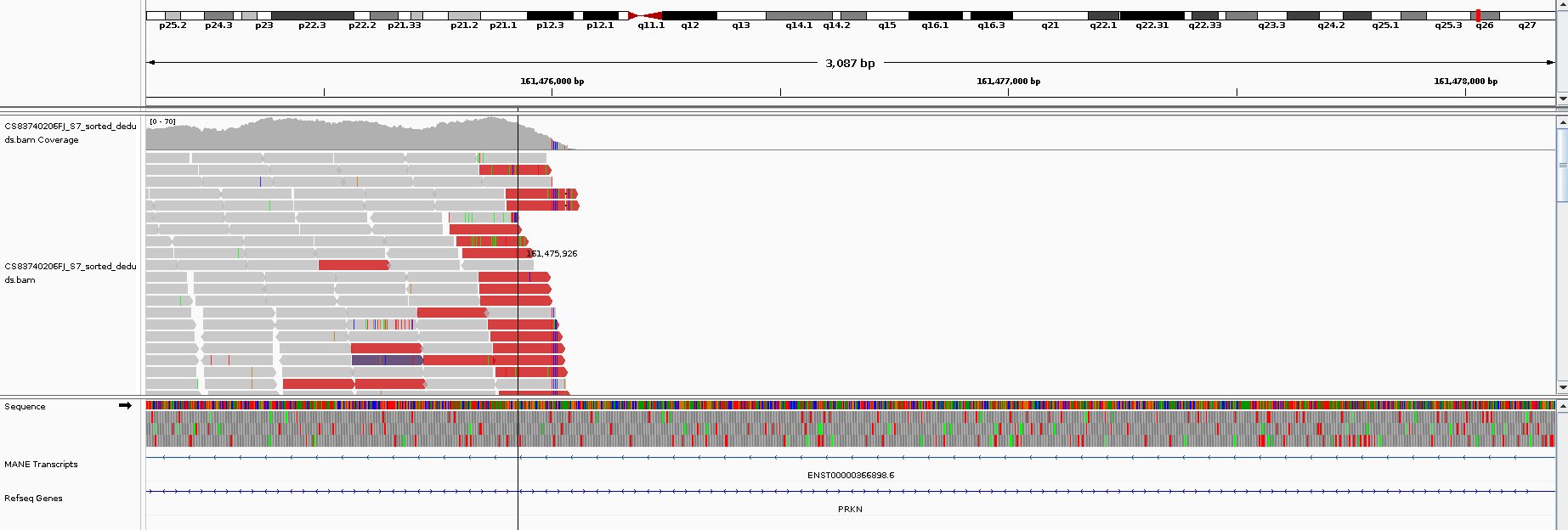

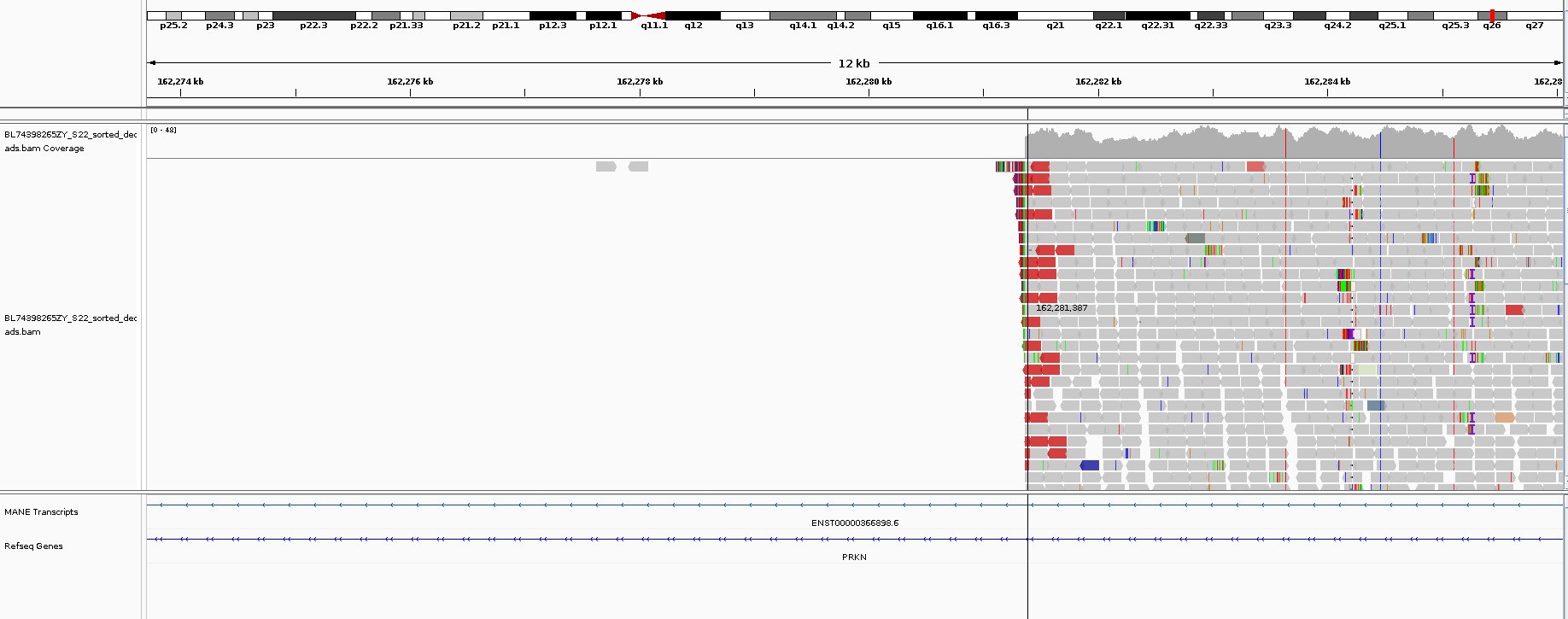

2. *GBA1* het deletion

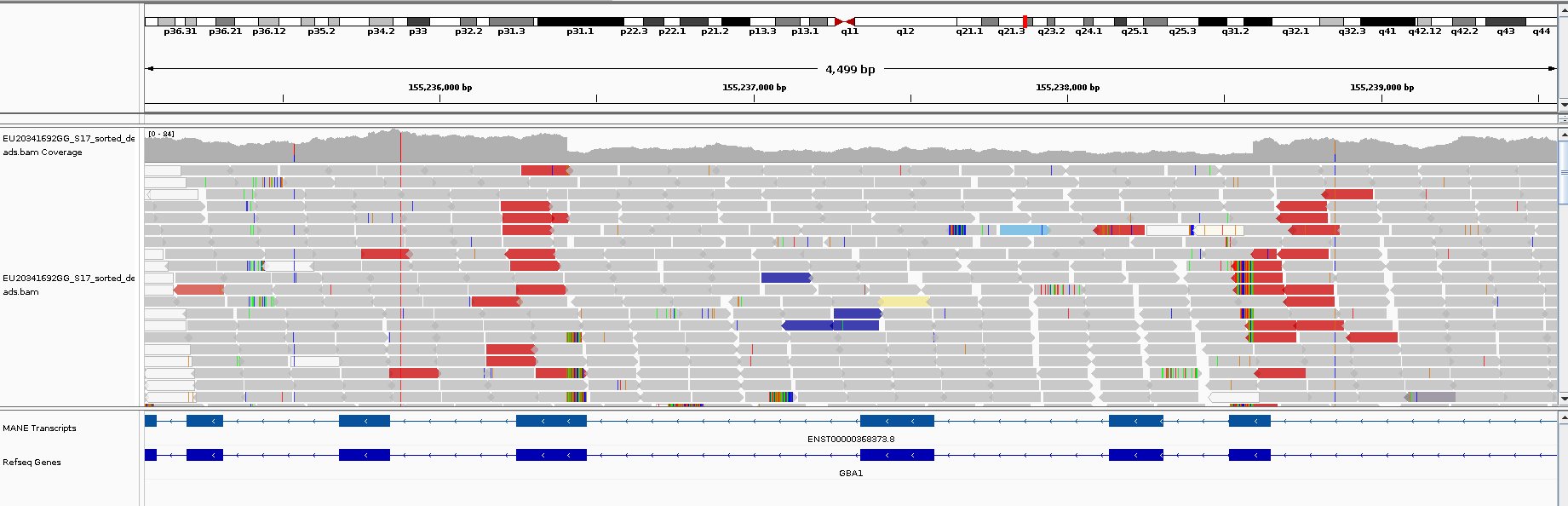

3. *CACNA1A4* het deletion

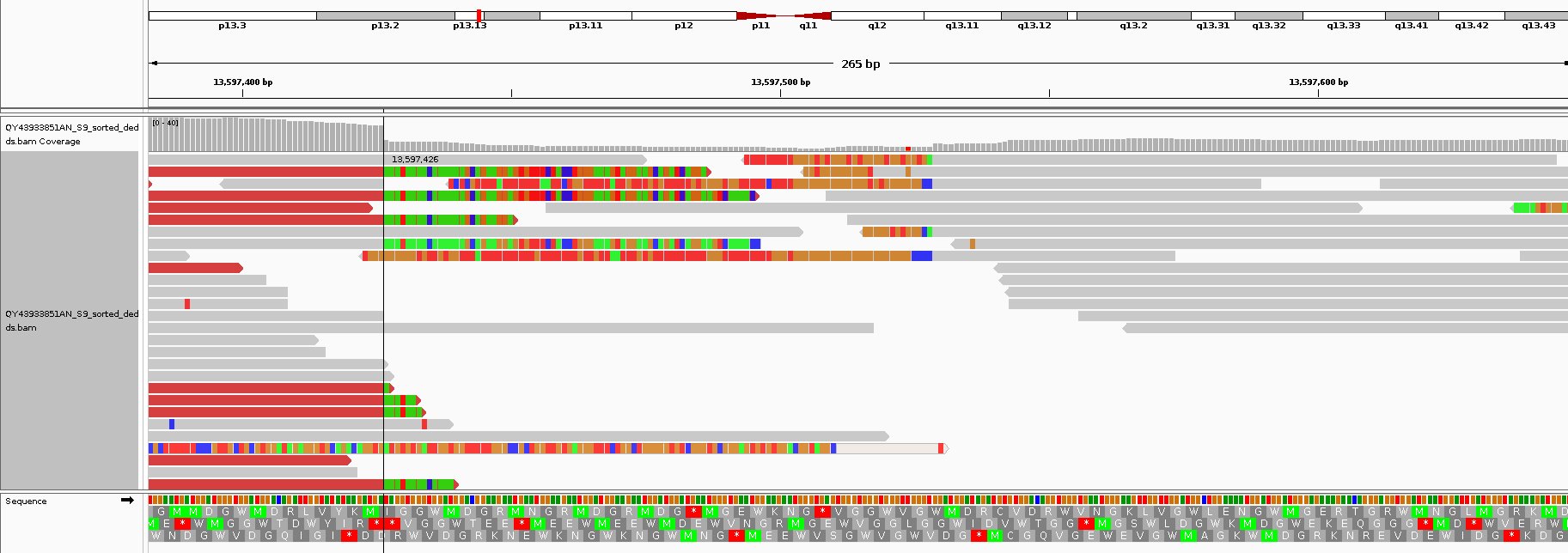

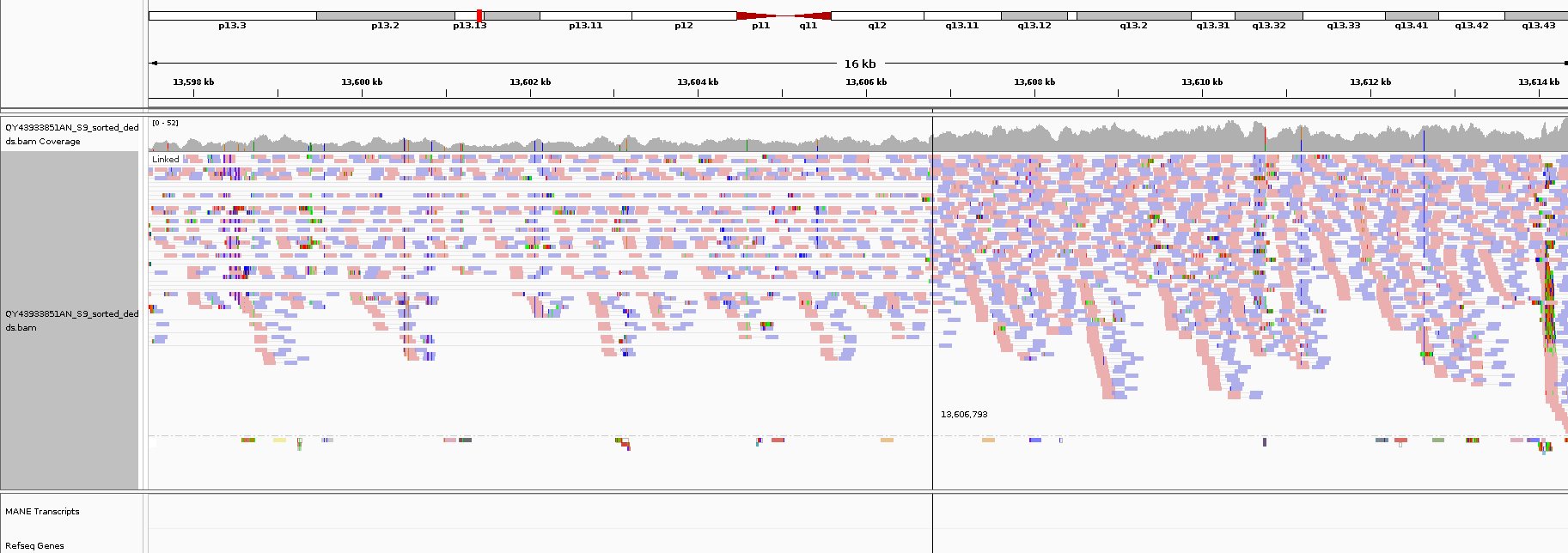

Supplementary Table Structure Description

| **Table Columns** |  |
| --- | --- |
| **Column name** | **Description** |
| Variant_ID | **Unique identifier for each variant (formatted as chromosome:position:ref:alt).** |
| Sample_ID | **Internal ID of the sample carrying the variant.** |
| Gene | **Gene associated with the variant.** |
| Inheritance | **Mode of inheritance (e.g., AR, AD, X-linked).** |
| Type | **Variant type (e.g., missense, nonsense, splice-site, deletion).** |
| Genotype | **Zygosity of the variant in the sample (e.g., het/hom).** |
| Consensus_Genotype | **Consensus genotype derived from all five SV callers used for structural variant detection.** |
| Novel/Known | **Indicates whether the variant is novel or previously reported in databases such as ClinVar or gnomAD.** |
| ACMG | **Classification based on ACMG guidelines (P = Pathogenic, LP = Likely Pathogenic, VUS = Variant of Uncertain Significance, LB = Likely Benign, B = Benign).** |
| Evidences | **Supporting lines of evidence used for ACMG classification.** |
| FamilyHistory | **Presence or absence of family history of Parkinson’s disease (PD).** |
| Freq | **Frequency within the study cohort.** |
| Control_count | **Number of occurrences in control samples.** |
| Phenotype | **Clinical category of the sample (e.g., YOPD, LOPD, HC).** |
| Canonical | **Indicates whether the variant affects the canonical transcript.** |
| Age Range | **Current range of age of the participant.** |
| Gender | **Participant sex (M or F).** |
| AAChange.ensGene | **Amino acid change annotation based on Ensembl transcript.** |
| AC | **Allele count observed in the dataset.** |
| AF | **Allele frequency observed in the dataset.** |
| AN | **Total number of alleles analyzed.** |
| CADD_phred | **CADD Phred-like score indicating predicted deleteriousness.** |
| CLNDISDB | **ClinVar disease database identifier(s).** |
| CLNDN | **ClinVar disease name(s).** |
| CLNREVSTAT | **ClinVar review status.** |
| CLNSIG | **ClinVar clinical significance.** |
| ClinPred_pred | **ClinPred prediction (D = Deleterious, T = Tolerated).** |
| Polyphen2_HVAR_pred | **PolyPhen-2 (HumVar) prediction (D = Probably damaging, P = Possibly damaging, B = Benign).** |
| Polyphen2_HVAR_rankscore | **PolyPhen-2 (HumVar) rank score.** |
| REVEL | **REVEL score for missense variant pathogenicity prediction.** |
| REVEL_rankscore | **Normalized rank score of REVEL.** |
| SIFT4G_converted_rankscore | **Normalized SIFT4G score.** |
| SIFT4G_pred | **SIFT4G prediction (D = Deleterious, T = Tolerated).** |
| AlphaMissense_pred | **AlphaMissense prediction (e.g., Pathogenic, Benign, Uncertain).** |
| AlphaMissense_pathogenicity_score | **Continuous pathogenicity score from AlphaMissense (0–1, higher = more pathogenic).** |
| gnomad40_genome_AF | **Allele frequency in gnomAD v4.0 (genome dataset).** |
| gnomad40_genome_AF_XX | **Allele frequency in female samples.** |
| gnomad40_genome_AF_XY | **Allele frequency in male samples.** |
| gnomad40_genome_AF_afr | **Allele frequency in African/African-American population.** |
| gnomad40_genome_AF_ami | **Allele frequency in Amish population.** |
| gnomad40_genome_AF_amr | **Allele frequency in Latino/Admixed American population.** |
| gnomad40_genome_AF_asj | **Allele frequency in Ashkenazi Jewish population.** |
| gnomad40_genome_AF_eas | **Allele frequency in East Asian population.** |
| gnomad40_genome_AF_fin | **Allele frequency in Finnish population.** |
| gnomad40_genome_AF_grpmax | **Highest observed allele frequency across all gnomAD populations.** |
| gnomad40_genome_AF_mid | **Allele frequency in Middle Eastern population.** |
| gnomad40_genome_AF_nfe | **Allele frequency in Non-Finnish European population.** |
| gnomad40_genome_AF_raw | **Raw allele frequency reported in gnomAD.** |
| gnomad40_genome_AF_remaining | **Allele frequency in unassigned or other populations.** |
| gnomad40_genome_AF_sas | **Allele frequency in South Asian population.** |
